# MRI-Derived Fat Depots and Cardiometabolic Pathways Underlying Heart Failure Risk: A Mendelian Randomization Study

**DOI:** 10.64898/2026.07.06.26357230

**Authors:** Pranav Sharma, Michael G. Levin

## Abstract

**Background:** Obesity is a major modifiable risk factor for heart failure (HF), but body mass index (BMI) does not distinguish biologically distinct fat depots. Whether imaging-derived adipose tissue depots capture specific cardiometabolic pathways underlying HF risk beyond conventional anthropometric measures remains uncertain.

**Methods:** We performed two-sample Mendelian randomization (MR) and multivariable MR mediation analyses using published genome-wide association study summary statistics. General adiposity traits included BMI, waist-to-hip ratio (WHR), and WHR adjusted for BMI from GIANT and UK Biobank meta-analyses of up to 694,649 individuals. MRI-derived visceral adipose tissue (VAT), abdominal subcutaneous adipose tissue (ASAT), and gluteofemoral adipose tissue (GFAT) were derived from 38,965 UK Biobank participants. HF outcome data were obtained from the HERMES consortium, including 1,946,349 individuals and 153,174 HF cases. Cardiometabolic mediators included type 2 diabetes (T2D), systolic blood pressure (SBP), LDL cholesterol, HDL cholesterol, and triglycerides. Primary analyses used inverse-variance weighted MR, with sensitivity analyses and directionality testing. Mediation was estimated using joint multivariable MR conditioning on significant cardiometabolic mediators.

**Results:** Among MRI-derived adipose depots, ASAT was the only trait significantly associated with HF risk (odds ratio [OR] 1.64 per 1-SD increase; 95% CI 1.40-1.93; FDR q<0.001). VAT showed a positive but imprecise association (OR 1.38; 95% CI 0.87-2.20), and GFAT was not associated with HF. Among general adiposity measures, BMI (OR 1.65; 95% CI 1.58-1.71) and WHR (OR 1.30; 95% CI 1.23-1.38) were robustly associated with HF, whereas WHR adjusted for BMI was not. ASAT was significantly associated with T2D, SBP, HDL cholesterol, and triglycerides, but not LDL cholesterol. In joint multivariable MR, 67.9% of ASAT’s HF effect was mediated through T2D, SBP, HDL cholesterol, and triglycerides (95% CI 49.2-86.8%). In contrast, BMI demonstrated only 8.5% mediation (95% CI -7.4 to 24.4%), and WHR showed non-significant mediation of 36.7% (95% CI -8.3 to 81.7%).

**Conclusions:** MRI-derived abdominal subcutaneous adipose tissue captures a biologically coherent cardiometabolic signal underlying HF risk that is diluted by conventional anthropometric measures. ASAT may represent an imaging biomarker of metabolic syndrome-mediated HF risk and could support more precise risk stratification and mechanistically targeted prevention in HF.

---

Obesity is a major modifiable risk factor for heart failure (HF), contributing to incident disease through both hemodynamic and metabolic mechanisms.^1^ However, body mass index (BMI), the common clinical measure to classify excess fat, captures overall adiposity without distinguishing metabolically distinct fat depots based on anatomic distribution.^2^

Large-scale MRI phenotyping in the UK Biobank has enabled characterization of discrete fat depots, revealing distinct genetic contributors.^3^ Mendelian randomization (MR) uses genetic variation as a natural experiment to investigate potentially causal relationships between modifiable risk factors and health outcomes.^4^ We applied this approach to evaluate whether MRI-derived fat depots better capture cardiometabolic pathways linked to HF compared to general adiposity measures.

We performed two-sample univariable MR and multivariable MR (MVMR) mediation analyses using published genome-wide association study (GWAS) data. We first tested the association of adiposity measures with HF. Next, we explored the effects of significant adiposity measures on cardiometabolic mediators. Finally, we performed MVMR to quantify the proportion of each fat trait’s HF effect operating through these pathways.

General adiposity traits (BMI, waist-to-hip ratio [WHR], and WHRadjBMI) were derived from GIANT+UK Biobank meta-analyses of up to 694,649 individuals.^5^ MRI-derived fat depot traits (visceral [VAT], abdominal subcutaneous [ASAT], and gluteofemoral [GFAT] adipose tissue) were derived from 38,965 UK Biobank participants.^3^ HF outcome data were obtained from the HERMES consortium (1,946,349 individuals; 153,174 cases).^6^ Cardiometabolic mediators included type 2 diabetes (T2D), systolic blood pressure (SBP), LDL cholesterol, HDL cholesterol, and triglycerides (TG), each from their respective largest GWAS.^7^

Genetic instruments were selected at genome-wide significance (P<5×10^−8^) with LD clumping (r^2^<0.001, 10,000-kb window). Primary analyses used inverse-variance weighted MR. Sensitivity analyses included weighted median, weighted mode, and MR-Egger regression. Steiger directionality testing and bidirectional MR were performed to assess potential reverse causation. False discovery rate (FDR) correction was applied using the Benjamini-Hochberg method (q<0.05). Mediation was estimated using the difference-of-coefficients method from joint MVMR conditioning simultaneously on all significant mediators, with 95% confidence intervals derived using the delta method. Analyses were performed in R using the TwoSampleMR package (https://mrcieu.github.io/TwoSampleMR).

Among MRI-derived adipose depots, ASAT was the only trait significantly associated with HF (OR 1.64 per 1-SD; 95% CI 1.40–1.93; q<0.001; Table 1). VAT showed a positive but imprecise association (OR 1.38; 95% CI 0.87–2.20; q=0.87), reflecting its small instrument set, and GFAT was null (OR 1.06; q=0.63). Among general adiposity measures, both BMI (OR 1.65; 95% CI 1.58–1.71) and WHR (OR 1.30; 95% CI 1.23–1.38) were robustly associated with HF; WHRadjBMI was not (OR 1.00; q=0.97). Sensitivity analyses remained directionally consistent and there was no evidence of reverse causation.

ASAT was significantly associated with four cardiometabolic mediators: T2D (OR 3.70; 95% CI 2.12–6.47), SBP (β=0.130, 95% CI 0.09–0.17), HDL (β=−0.144, 95% CI −0.26 to −0.03), and TG (β=0.158, 95% CI 0.10–0.22). LDL was not significantly associated with any adiposity measure. BMI and WHR showed significant associations with T2D, SBP, HDL, and TG (all FDR q<0.05). These mediators were independently associated with HF in univariable MR (q<0.05).

In joint MVMR conditioning simultaneously on T2D, SBP, HDL, and TG, 67.9% (95% CI: 49.2–86.8%) of ASAT’s HF effect was mediated through these classical cardiometabolic pathways; the only fat trait to achieve statistically significant mediation (Table 1, Figure 1). BMI demonstrated only 8.5% mediation (95% CI: −7.4 to 24.4%). WHR showed intermediate but non-significant mediation (36.7%, 95% CI: −8.3 to 81.7%). SBP was the dominant mediating pathway in joint models for all three fat traits.

**Figure 1.**
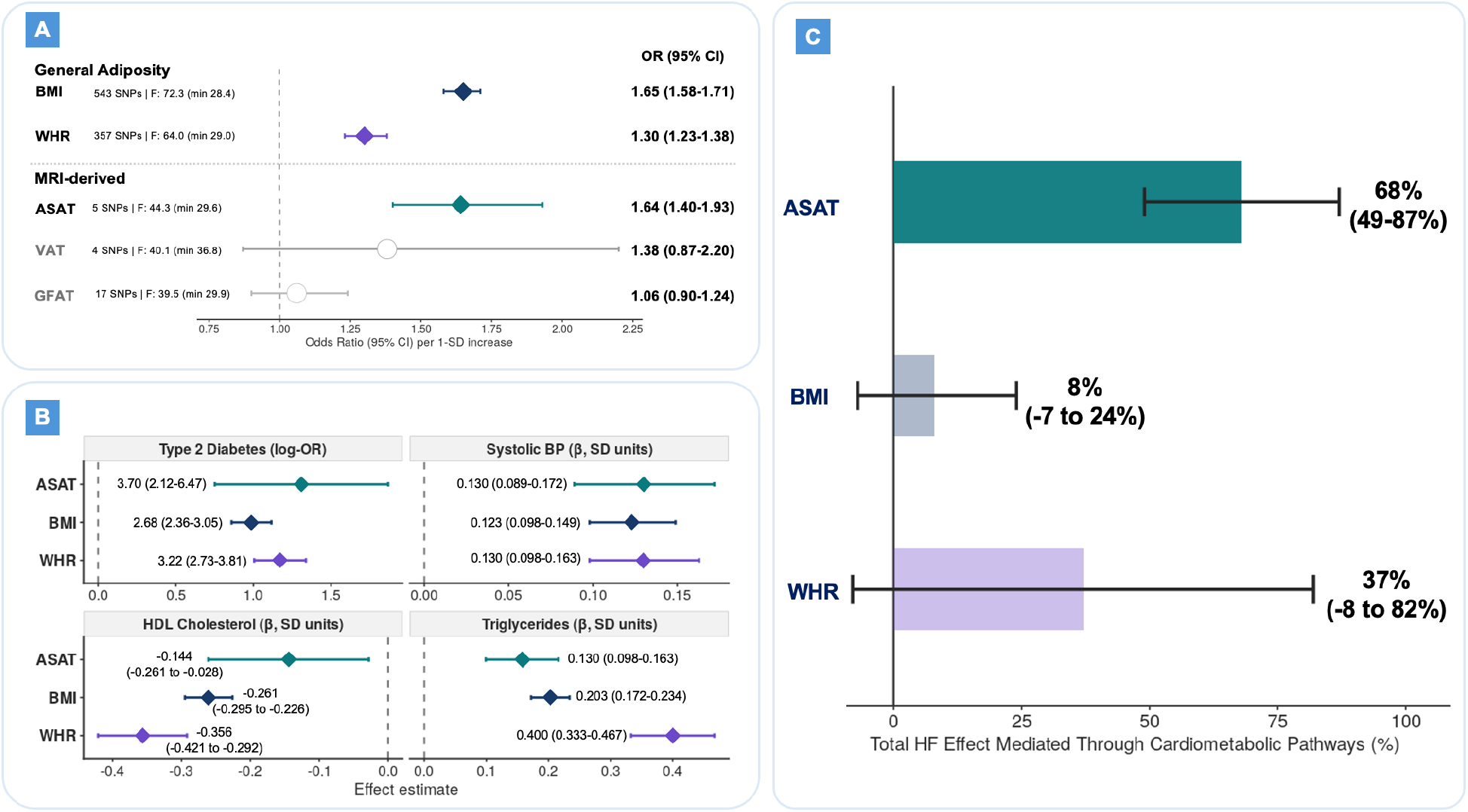
MRI-derived fat depot measures capture cardiometabolic pathways underlying heart failure risk. (A) Inverse-variance weighted Mendelian randomization estimates for the association of genetically predicted fat traits with heart failure risk, reported as odds ratios per 1-SD increase. Filled diamonds indicate associations significant at FDR q<0.05. Number of SNPs, mean F-statistic, and minimum F-statistic are shown next to trait labels. (B) Associations of genetically predicted ASAT, BMI, and WHR with cardiometabolic risk factors implicated in heart failure pathogenesis. Type 2 diabetes is shown as an odds ratio; SBP, HDL cholesterol, and triglycerides are shown as beta estimates. (C) Proportion of each fat trait’s total heart failure effect mediated through cardiometabolic pathways in joint multivariable Mendelian randomization conditioning simultaneously on type 2 diabetes, systolic blood pressure, HDL cholesterol, and triglycerides. Error bars represent 95% CIs estimated using the delta method. ASAT demonstrated significant mediation through these pathways, whereas BMI and WHR did not.

Our results demonstrate that ASAT exerts a significant effect on HF risk, and its effect operates predominantly through established cardiometabolic mechanisms. The mediating pathways highlighted in our analysis collectively reflect core features of metabolic syndrome, suggesting that ASAT may function as an imaging biomarker of metabolic syndrome-mediated HF risk. In contrast, conventional measures of excess adiposity, including BMI and WHR, remain largely unexplained by these same pathways, suggesting they may capture risk mediated by other processes, which may include hemodynamic loading, neurohormonal activation, adipokine signaling, and systemic inflammation.^8,9^ These findings suggest that enriching clinical trials of emerging cardiometabolic therapies, including GLP-1s, SGLT2s, and lipid pathways, for individuals with ASAT-driven HF risk may be a promising area for future research, particularly given the growing number of therapeutic targets in this space.^10,11^ These findings complement recent evidence from a large-scale MR study demonstrating that genetically predicted BMI is associated with increased mortality and cardiovascular hospitalization across the ejection fraction spectrum in ∼50,000 patients with established HF, underscoring the importance of understanding the mechanistic basis through which adiposity confers HF risk.^12^

These findings are also timely in the context of rapidly expanding AI-derived imaging phenotype initiatives. The UK Biobank, as used in this study, has established the feasibility of MRI-derived fat depot quantification at scale, and deep learning pipelines now segment ASAT, VAT, and GFAT with high accuracy from both MRI and more clinically available routine CT imaging.^13,14^ Together, these advances suggest that ASAT quantification from imaging may better capture the metabolic syndrome-driven HF risk currently approximated by BMI in clinical practice, enabling more precise risk stratification, targeted prevention, and treatment in HF.

This study must be interpreted in the context of its limitations. Firstly, MRI-derived exposure GWASs are derived from the UK Biobank imaging subset (n=38,965), limiting instrument strength and statistical power relative to the general adiposity GWASs (n=694,649). Although partial sample overlap may exist between exposure and outcome GWASs, bias is unlikely given the comparatively small imaging subset.^15^ Second, GWAS data are predominantly of European-ancestry individuals, which may limit generalizability. Third, MR assumes lifelong exposure to risk factors of interest and may not reflect short-term intervention efforts. Finally, we did not explore sub-types of heart failure or sex-stratify these analyses, warranting further research to clarify these effects in specific sub-populations.

MRI-derived fat depot measures capture a biologically coherent cardiometabolic signal underlying HF risk that anthropometric traits dilute. ASAT may represent an imaging biomarker of metabolic syndrome-mediated HF risk. ASAT quantification from routine imaging may offer a path toward precision risk stratification and mechanistically targeted prevention in HF.

## Abbreviations

ASAT: abdominal subcutaneous adipose tissue
BMI: body mass index
CI: confidence interval
FDR: false discovery rate
GFAT: gluteofemoral adipose tissue
HDL: high-density lipoprotein
HF: heart failure
IVW: inverse-variance weighted
MR: Mendelian randomization
MVMR: multivariable Mendelian randomization
SBP: systolic blood pressure
T2D: type 2 diabetes
VAT: visceral adipose tissue
WHR: waist-to-hip ratio

## Ethical approval

This study used publicly available, de-identified GWAS summary statistics. All contributing studies obtained relevant ethical approval and participant informed consent. The present analysis did not involve individual-level human participant data and did not require additional institutional review board approval.

## Data availability

This study used publicly available GWAS summary statistics. Source datasets are available from the cited genome-wide association studies and the MRC IEU OpenGWAS platform. No new individual-level data were generated.

## Author contributions

P.S. and M.G.L. conceived the study. P.S. performed the analyses and drafted the manuscript. M.G.L. supervised the study and critically revised the manuscript. Both authors interpreted the data, approved the final manuscript, and agree to be accountable for the work.

